# Incidence and outcomes of unstable angina in patients with low high-sensitivity cardiac troponin I values – A substudy of the RACE-IT trial

**DOI:** 10.1101/2025.08.05.25333080

**Authors:** Raef Fadel, Joseph Miller, Bernard Cook, Felix Nguyen, Mohammad Alqarqaz, Brittany Fuller, Mir Basir, Tiberio Frisoli, Pedro Villablanca, Ahmad Jabri, Khaldoon Alaswad, Akshay Khandelwal, Natesh Lingam, Brian O’Neill, Henry Kim, Pedro Engel Gonzalez, Elizabeth Pielsticker, Gerald Koenig, Seth Krupp, Nicholas L Mills, Simon Mahler, Phillip Levy, Benjamin Brennan, Shane Bole, Sachin Parikh, Khaled Nour, Michael Hudson, Bryan Zweig, Omr Abuzahrieh, Chaun Gondolfo, James McCord

**Author notes:** Corresponding Author: Raef Fadel DO, Henry Ford Hospital, 2799 W Grand Blvd, K-14 Cardiovascular Medicine, Detroit, MI, 48202. **Funding:** None. **Disclosures:** No relevant disclosures.

## Abstract

**Background:** Unstable angina has become an exceedingly rare diagnosis in the era of high-sensitivity cardiac troponin (hs-cTn). We sought to identify the incidence of unstable angina and characterize patients with low hs-cTn in Emergency Departments (EDs).

**Methods:** A prespecified secondary analysis of the **R**apid **A**cute **C**oronary Syndrome **E**xclusion using high-sensitivity **I** cardiac **T**roponin (RACE-IT) trial was conducted. RACE-IT was a stepped-wedge randomized trial comparing two rule-out protocols (0/1- and 0/3-hour) for myocardial infarction (MI) in nine EDs from July 2020 to April 2021. All patients had hs-cTnI (Beckman Coulter) concentrations below or equal to the 99^th^ percentile upper reference limit of 18 ng/L. The primary outcome was unstable angina, based on the **ISCHEMIA** trial definition, which required electrocardiographic changes or findings at coronary angiography (angiographic evidence of plaque rupture or thrombus).

**Results:** Of the 32,608 patients in the trial, 60 patients (0.2%) met the definition of unstable angina of which 46 (77%) had obstructive disease at coronary angiography and 17 (28%) had an ischemic electrocardiogram. Coronary revascularization was performed in 45 (75%) patients and 7 (12%) had left main or 3-vessel coronary artery disease. There were 7 (12%) patients with non-obstructive coronary artery disease, and 7 (12%) who had angiographically unremarkable coronary arteries. Patients with unstable angina were older (p=0.032), more likely to be male (p=0.012), with a higher prevalence of hypertension (p<0.001), known coronary artery disease (p<0.001), peripheral vascular disease (p=0.030), and a higher serum creatinine (p=0.043). At 30 days, two patients had a type 1 MI and there were no deaths.

**Conclusion:** Unstable angina was diagnosed in 1 in 500 patients with a low hs-cTnI value at presentation to the ED and these patients have an excellent prognosis at 30 days. These patients tend to not have high-risk anatomy and 1 in 4 have non-obstructive coronary artery disease or angiographically unremarkable coronary arteries.

## Background

Utilization of high-sensitivity cardiac troponin (hs-cTn) for the rapid diagnosis or exclusion of myocardial infarction (MI) in the emergency department (ED) is the standard of care for the management of patients with possible MI and is recommended by guidelines ^1–2^. Patients with symptoms of myocardial ischemia at risk for major adverse cardiac events (MACE) who do not meet the criteria for MI, are often diagnosed as having unstable angina ^3,4^, but the diagnosis is more subjective and has been variably applied in clinical practice ^5^. This is primarily due to the reliance on the clinical history for the diagnosis, despite the fact that symptoms of myocardial ischemia are heterogeneous ^3^.

The true incidence of unstable angina is unclear, especially since the adoption of cTn testing and particularly with the more recent use of hs-cTn assays. Many patients with unstable angina in prior studies utilizing creatinine kinase (CK-MB) would have been reclassified as MI if a more sensitive cTn assay had been used ^5^. The American College of Cardiology and American Heart Association guidelines for management of patients with acute coronary syndrome recognize that close to a third of patients who have previously been diagnosed with unstable angina without CK-MB elevation have cTn elevation and would be reclassified as MI ^6^. With the introduction of hs-cTn assays, the incidence of unstable angina has been reduced further ^7^.

The RACE-IT (**R**apid **A**cute **C**oronary Syndrome **E**xclusion using high-sensitivity cardiac **I T**roponin) trial recently compared an accelerated protocol (AP) over 1 hour to a standard care (SC) protocol over 3 hours using a hs-cTnI assay in the diagnosis and management of patients with possible MI ^8^. In this prespecified secondary analysis of the trial we sought to evaluate the incidence of unstable angina and characterize these patients.

## Methods

### Study Design

The RACE-IT trial was a stepped-wedge, randomized controlled trial of consecutive patients evaluated for possible MI across the nine hospitals from Henry Ford Health in Detroit, Michigan from July 2020 to April 2021. Details of this study have previously been published ^8,9^. Patients were included if they had hs-cTnI measured using the Access Beckman Coulter high-sensitivity assay and an electrocardiogram (ECG) was obtained in the ED. Exclusion criteria included age less than 18 years, ST-segment elevation MI, trauma, transfers from other facilities, residence outside of Michigan, resident in hospice, and hs-cTnI above the 99th percentile upper reference limit (URL) of 18 ng/L on testing within the first 3 hours from presentation to the ED. RACE-IT compared a 1-hour protocol where all hs-cTnI values >4ng/L were reported to a 3-hour protocol where only values ≥18 ng/L were reported. The assay has a limit of detection of 0.3 ng/L and a limit of quantification of 0.8 ng/L ^10^.

In this analysis, our aim was to identify and characterize patients with unstable angina and to determine their Major adverse cardiac event (MACE) rate at 30 days. MACE was defined as subsequent MI or death from any cause at 30 days from presentation. All patients from the RACE-IT trial were included in this secondary analysis, which then stratified patients as having unstable angina or not.

### Definitions and Data Adjudication

Unstable angina was defined according to the criteria used in the ISCHEMIA trial.^11^ Unstable angina was considered in those with ischemic symptoms at rest, or an accelerating pattern that occurs with a lower activity threshold resulting in a visit to a healthcare facility, with cardiac troponin concentrations below the 99^th^ percentile URL, and at least one of the following: 1) new or worsening ST-segment or T-wave changes on a resting electrocardiogram consistent with myocardial ischemia, or 2) angiographic evidence of a ruptured plaque or thrombus in an epicardial coronary artery thought to be responsible for the ischemic symptoms or signs. Obstructive coronary artery disease was defined as a stenosis ≥70% in a major epicardial vessel, ≥50% in the left main stem, or evidence of flow limitation by physiologic testing including fractional flow reserve or instantaneous wave-free ratio.

Cases were adjudicated by interventionalist cardiologists, general cardiologists, or cardiology fellows. All patients that had coronary angiograms were reviewed by interventional cardiologists to determine unstable angina. Cases of patients that did not have a coronary angiogram but an elevation of hs-cTnI > 18 after admission, were reviewed by cardiology fellows to determine unstable angina. Outcomes at 30 days were identified by review of the electronic medical record (Epic Systems). An interventional cardiologist adjudicated MI in accordance with the universal definition of MI ^12^. Twelve institutions in Michigan use Epic and participate in a health information exchange. Death at 30 days was determined by review of the electronic medical record and by accessing the National Death Index. For deaths determined by the National Death Index, diagnostic codes consistent with a cardiac cause of death (120-125,146, 149) were used to classify cardiac death ^13^. All cases of cardiac death or MI at 30 days were reviewed by a general cardiologist to determine if the original presentation constituted unstable angina. Cardiology fellows also reviewed all the ECGs who were adjudicated to have unstable angina to see if they met the ISCHEMIA trial definition of ischemic ECG changes.

### Statistical analysis

Demographic and comorbid variables in US patients were compared to the corresponding variables in the non-unstable angina cohort using appropriate statistical tests for each variable type and sample size. All continuous variables were analyzed using the Wilcoxon Rank-Sum Test. Due to small sample size, differences in peripheral vascular disease were assessed using Fisher’s Exact Test – all other categorical or binary variables were analyzed using a Chi-Squared Test. All analysis was performed using R version 4.4.0.

## Results

### Baseline characteristics and details of presentation

Of the 32,608 patients enrolled in the RACE-IT trial, 60 patients (0.2%) met the definition of unstable angina of whom 37 (62%) were enrolled from the group evaluated using the advanced protocol and 23 (38%) in the group evaluated using the standard care protocol. Patients with unstable angina were older (p=0.032) and were more likely to be male (p=0.012). There was a higher prevalence of hypertension (p<0.001), previous coronary artery disease (p<0.001), and peripheral vascular disease (p=0.030) among patients with unstable angina. Additionally, patients with unstable angina had higher creatinine concentrations (p=0.043) with a trend toward a lower estimated glomerular filtration rate (p=0.067)(**Table 1**).

**Table 1.**
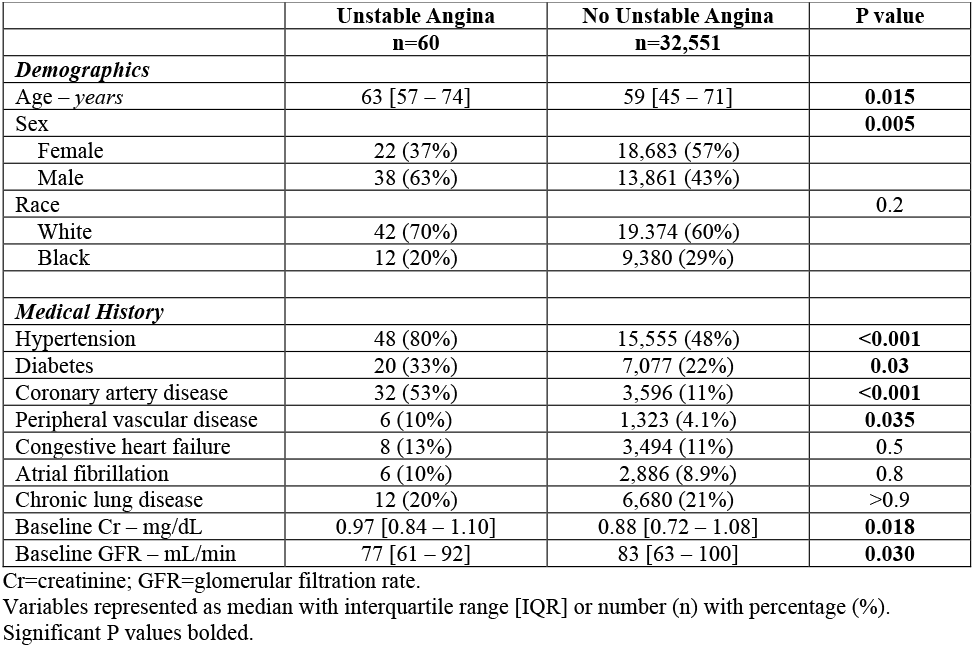
Baseline characteristics.

|  | Unstable Angina<br>n=60 | No Unstable Angina<br>n=32,551 | P value |
| --- | --- | --- | --- |
| ***Demographics*** |  |  |  |
| Age – years | 63 [57 – 74] | 59 [45 – 71] | **0.015** |
| Sex |  |  | **0.005** |
| Female | 22 (37%) | 18,683 (57%) |  |
| Male | 38 (63%) | 13,861 (43%) |  |
| Race |  |  | 0.2 |
| White | 42 (70%) | 19,374 (60%) |  |
| Black | 12 (20%) | 9,380 (29%) |  |
| ***Medical History*** |  |  |  |
| Hypertension | 48 (80%) | 15,555 (48%) | **<0.001** |
| Diabetes | 20 (33%) | 7,077 (22%) | **0.03** |
| Coronary artery disease | 32 (53%) | 3,596 (11%) | **<0.001** |
| Peripheral vascular disease | 6 (10%) | 1,323 (4.1%) | **0.035** |
| Congestive heart failure | 8 (13%) | 3,494 (11%) | 0.5 |
| Atrial fibrillation | 6 (10%) | 2,886 (8.9%) | 0.8 |
| Chronic lung disease | 12 (20%) | 6,680 (21%) | >0.9 |
| Baseline Cr – mg/dL | 0.97 [0.84 – 1.10] | 0.88 [0.72 – 1.08] | **0.018** |
| Baseline GFR – mL/min | 77 [61 – 92] | 83 [63 – 100] | **0.030** |

Cr=creatinine; GFR=glomerular filtration rate.
Variables represented as median with interquartile range [IQR] or number (n) with percentage (%).
Significant P values bolded.

### Diagnostic results in patients with unstable angina

In the overall study there were 400 coronary angiograms performed. All but 2 of the patients had their coronary angiograms during the index hospitalization, and all were performed within 30-days of index presentation. Results of the coronary angiograms of the unstable angina patients are shown **(Table 2)**. There were 46 (77%) patients that had obstructive disease, and all but 1 underwent a revascularization procedure (41 percutaneous interventions and 4 coronary artery bypass surgeries). Of note, only 7 (12%) had high-risk coronary disease (left main or 3-vessel CAD), 7 (12%) had non-obstructive disease, and 7(12%) had angiographically unremarkable coronary arteries. All unstable angina patients with angiographically unremarkable coronary arteries or non-obstructive disease were noted to have ischemic ECG changes.

**Table 2.**
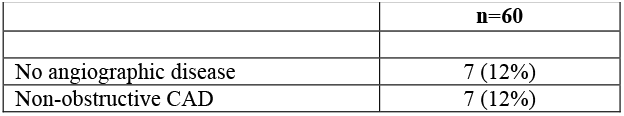
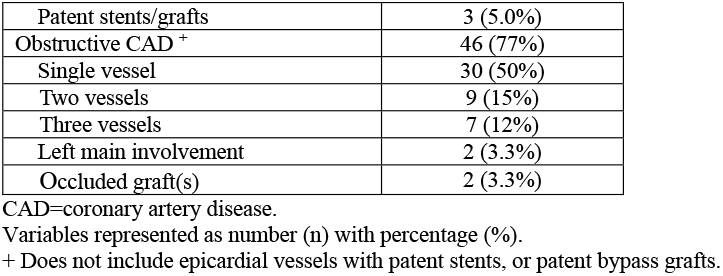
Coronary Angiography in Unstable Angina Patients.

On presentation, 17 (28%) of the patients classified as having unstable angina following angiographic review had evidence of myocardial ischemia on the electrocardiogram including T-wave inversion (25%) and/or ST-segment depression (13%) **(Table 3)**. Also, 12 patients (20%) in the unstable angina group had extremely low hs-cTnI values < 4ng/L.

**Table 3.**
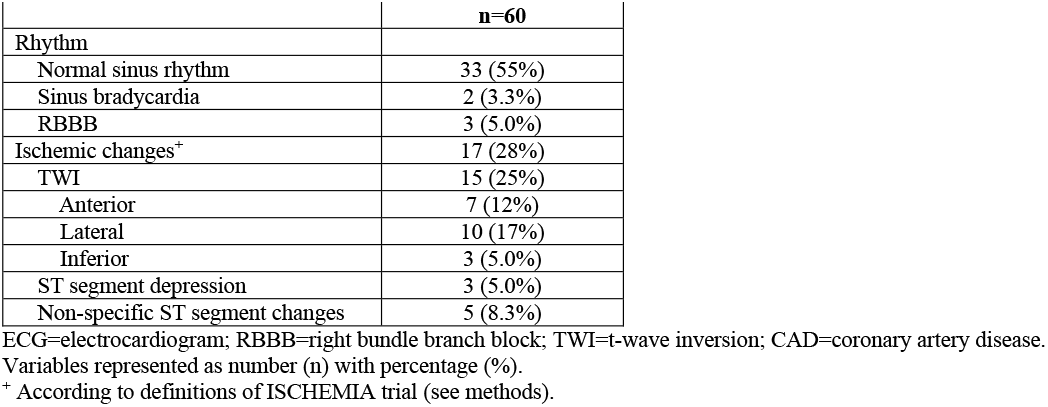
ECG results in Unstable Angina Patients.

### Outcomes

In patients without unstable angina, 26 (0.1%) experienced MI and 64 (0.3%) died of any cause at 30-days with 8 of these deaths attributed to a cardiac cause. There were no deaths at 30 days in patients classified as having unstable angina. However, there were 2 (3%) patients with a type 1 MI at 30 days. The first patient was an 86-year-old male with known coronary artery disease presented with episodic chest pain at rest without myocardial ischemia on the electrocardiogram in whom all hs-cTnI concentrations were <18 ng/L. He was advised to stay for further evaluation but he wanted to leave to follow-up with his cardiologist. He presented 5 days later with an inferior ST-segement elevation MI and underwent PCI to the right coronary artery. The second patient was a 75-year-old male with known coronary artery disease who presented with chest pain that was similar to the symptoms he experienced during a previous MI. He had no myocardial ischemia on the electrocardiogram and had serial hs-cTnI values of 6 and 7 ng/L. He had a nuclear stress test within in the past year that showed a small area of ischemia in the left circumflex distribution, and he was discharged home. He returned 2 weeks later with a diagnosis of non-ST-segment elevation MI. He underwent PCI to the right coronary artery.

## Discussion

In this prespecified secondary analysis of the RACE-IT trial we report the incidence of unstable angina in a large consecutive patient population evaluated for possible MI in whom serial hs-cTnI measurements were within the normal reference range at presentation. We report three major findings.

First, the incidence of unstable angina in patients with hs-cTnI values < 99^th^, using the definition from the ISCHEMIA trial, is very low, only 0.2% in our study. This is in line with a prior study which have shown the incidence of unstable angina decreases with the introduction of hs-cTn assays. Wilson et al reported similar findings when assessing patients enrolled in the Randomized Trial to Evaluate the Relative PROTECTion against Post-PCI Microvascular Dysfunction (PROTECT)-TIMI 30 trial. With use of a hs-cTnI assay with a detection limit down to 0.2 ng/L and 99^th^ percentile reference limit of 3.0 ng/mL, compared to prior standard commercially available assays at the time, 44% of patients previously diagnosed with unstable angina demonstrated troponin levels at presentation exceeding the 99^th^ percentile, increased to 82% of patients by the 8 hour mark ^14^.

Second, the prognosis at 30 days is excellent in patients with confirmed unstable angina. There were no patients who died and only 2 had a MI at 30 days. This could be explained by the low prevalence of high-risk coronary anatomy involving the left main stem or all 3 major epicardial coronary arteries. Other studies have reported similar findings. Gallone et al reported that in 240 patients with unstable angina in whom hs-cTnT concentrations were <99^th^ percentile there were no deaths at 30 days and only 0.8% had a MI ^15^. Similarly, Dakshi et al reported that 158 patients with unstable angina with hs-cTnT concentrations <99^th^ percentile had no deaths at 30 days and 3 (2%) MIs ^16^. While prior studies investigated the incidence of unstable angina when using an hs-cTnT assay, our study is the first to evaluate the incidence using an hs-cTnI. Evaluation of both hs-cTnI and hs-cTnT is important in this situation because some differences have been noted between these assays ^17^.

Third, a significant number of patients with unstable angina in this study had angiographically unremarkable or non-obstructive disease (14 patients, 23%). Although this may seem surprising other studies have shown similar results. In the study by Paiva et al, 47% patients with unstable angina had non-obstructive coronary disease on catheterization ^18^. Also, it is well-known that coronary vasospasm can present as unstable angina ^19^. In addition, it is now well-known that that 6%-8% of patients with MI have myocardial infarction with non-obstructive coronary arteries (MINOCA) ^20^. It is possible in the overall ACS arena that this is more common in unstable angina.

### Limitations

There are several limitations worth considering when analyzing the results of this study. First and foremost, this was a subgroup analysis of the main RACE-IT trial, and was not the main aim of the primary study. Also not all patients with ischemic ECGs were adjudicated. Only patients that went onto coronary angiography, had elevation of hs-cTnI > 18 ng/L after ED presentation, or type 1 MI or cardiac death at 30 days were evaluated. Also, treatment of patients presenting to the ED enrolled in the primary RACE-IT trial was left to the discretion of the treating physicians, and therefore as a subgroup analysis this study had no control over who went to catheterization and who did not. A significant amount of clinical decision making factors into the treatment of such patients, and this is acknowledged by the research team.

## Conclusion

Unstable angina is rare in patients with a low hs-cTnI values at presentation to the ED and these patients have an excellent 30-day prognosis. These patients tend to not have high-risk anatomy and a significant number have non-obstructive disease or angiographically unremarkable coronary arteries.

## Data Availability

All data is property of the host institution and will not be made readily available, or will be shared upon request.

## Abbreviations List

CK-MB: creatinine kinase
CAD: coronary artery disease
ECG: electrocardiogram
ED: emergency department
Hs-cTnI: high-sensitivity cardiac troponin I
MACE: major adverse cardiovascular event
MI: myocardial infarction

---

**Figure 1.**
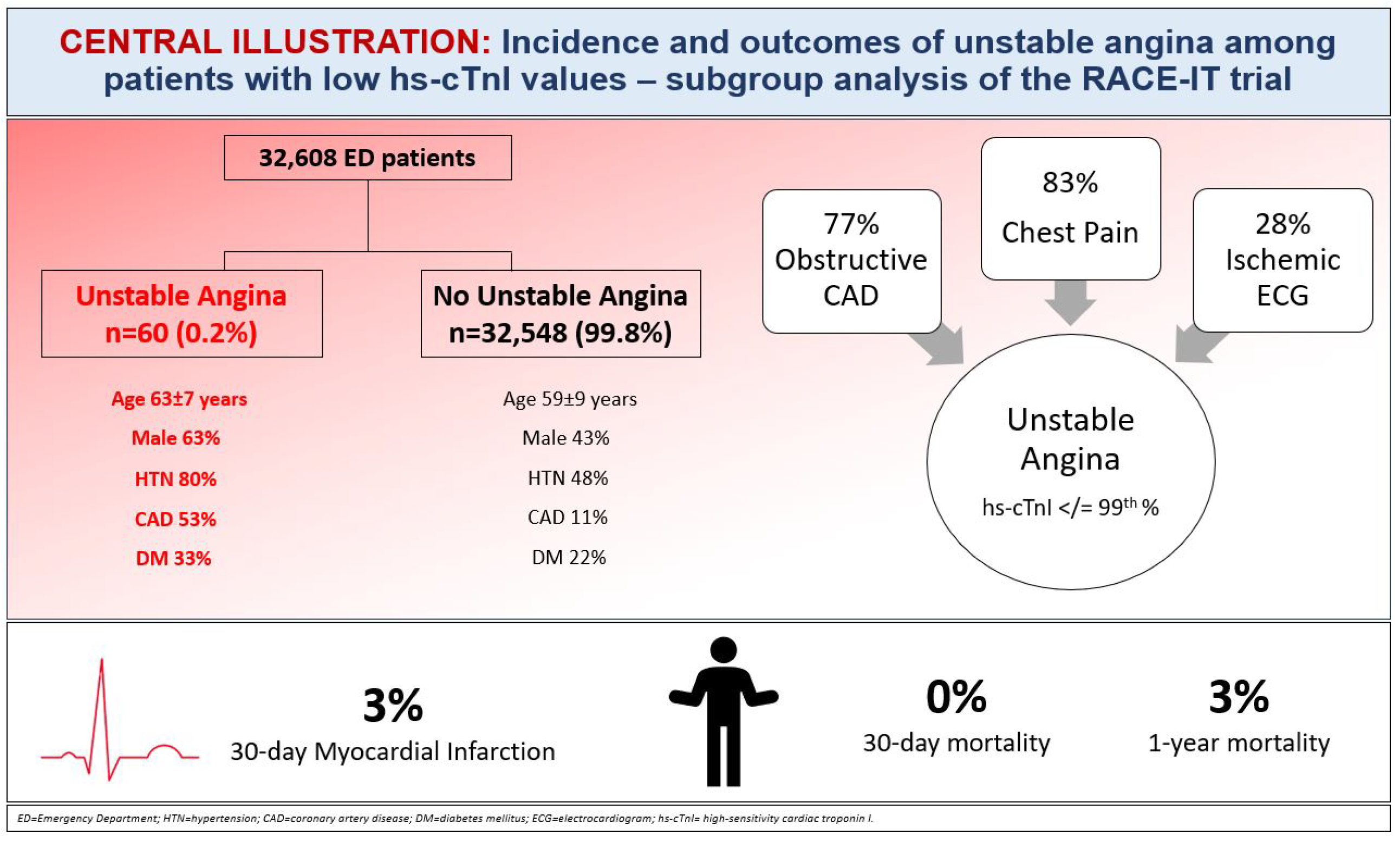
Central illustration. *ED=Emergency Department; HTN=hypertension; CAD=coronary artery disease; DM=diabetes mellitus; ECG=electrocardiogram; hs-cTnI= high-sensitivity cardiac troponin I*.

## Notes

### Competing Interest Statement

The authors have declared no competing interest.

### Funding Statement

Non-funded study.

### Author Declarations

Henry Ford Hospital IRB

